# Baseline hypocapnia is associated with intubation in COVID-19 diagnosed patients

**DOI:** 10.1101/2021.11.19.21266581

**Authors:** Athanasios Gounidis, Alexandros P. Evangeliou, Christina Kloura, Evangelia Manganari, Christiana Parisi, Michalis Kourtidis, Georgios Kotronis, Martha Apostolopoulou, Fani Apostolidou-Kiouti

## Abstract

**Introduction:** Hypocapnia may be one of the several factors predefining the need for intubation of patients needing hospitalization for COVID-19 pneumonia.

**Methods:** A retrospective evaluation of patient files hospitalized for COVID-19 pneumonia from October 2020 until January 2021. Univariate and multivariate regression was used, as well as a multinomial regression to account for multiple endpoints (discharge, intubation, death).

**Results:** Hypocapnia was strongly associated with intubation (OR: 0.86, 95% CI: 0.76, 0.97). Additionally, last pCO2 (OR: 1.08, 95% CI: 1.01, 1.16), baseline FiO2 (OR: 1.05, 95% CI: 1.03, 1.07) as well as last FiO2 (OR: 1.21, 95% CI: 1.11, 1.46), total severity score on admission (OR: 1.18, 95% CI: 1.03, 1.37) and last pO2 (OR: 0.89, 95% CI: 0.85, 0.92) were found to have a significant impact on intubation. Incorporation of deceased patients withheld the negative association with pCO2 levels (OR: 0.88, 95% CI: 0.78, 0.98).

**Conclusion:** The dissociation between respiratory failure and a clinically comfortable patient is partly due to decreased carbon dioxide levels and clinicians should bare it in mind when handling patients with COVID-19 pneumonia. Hypocapnia seems to be a determinant factor of intubation in patients with COVID-19 pneumonia in this study.

## INTRODUCTION

The severe acute respiratory syndrome (Sars-CoV-2) pneumonia was first diagnosed in December of 2019 in China and quickly grew to a pandemic with significant impact on health systems worldwide. A wide spectrum of clinical severity has been reported, ranging from asymptomatic to critically ill patients^1, 2^. Recently published systematic reviews reported that the most frequent symptoms of these patients are fever and cough followed by shortness of breath and myalgia^3, 4^. A major clinical characteristic, complicating patient care, is the paradoxical lack of dyspnea despite extremely low hemoglobin saturation and partial pressure of oxygen (pO_2_) in arterial blood; two thirds of coronavirus disease 2019 (COVID-19) hospitalized patients had not complained of shortness of breath on admission, despite present signs of pneumonia in computed tomography chest scan ^5, 6^ and low pO_2_ in arterial blood gases (ABGs). Hypoxemia without respiratory distress has previously been associated with poor outcome in patients developing acute respiratory distress syndrome (ARDS), with the majority of patients being intubated or died ^7^. The pathophysiological mechanism for hypoxia upon admission has yet to be determined, while association of hypoxia with the thrombosis of pulmonary vessels has also been described^8^. This association could be due to several mechanisms including ventilation/ perfusion (V/Q) mismatch, thrombotic events, SARS-CoV-2 specific effect on chemoreceptors, altered diffusion capacity or loss of vasoadaptive mechanisms ^9^.

Happy or silent hypoxia seems to have a significant pathophysiological role in the clinical presentation of patients with COVID-19 pneumonia^10^; a state in which low pO2 along with normal or low partial pressure of carbon dioxide (pCO2) and normal or increased alveolar ventilation are not perceived as respiratory discomfort. Since hypocapnia has already been related with poorer outcomes in patients requiring invasive mechanical ventilation (IMV) ^11, 12^, this study was conducted to investigate a possible association between carbon dioxide levels in the early stages of disease and progression to intubation.

## DESIGN – METHODS

### Study design

This is retrospective cohort study which was conducted in our centre between October 2020 and January 2021. It is a referral hospital assigned to hospitalize patients undergoing reallocation from other centres due to full occupancy. All the medical reports from patients were collected and data from medical records were further analyzed.

### Participants

Consecutive patients older than 18 years of age, with or without previously diagnosed pulmonary disease, irrespective of smoking status, race or gender, diagnosed with COVID-19 pneumonia confirmed with nasopharyngeal swab test (polymerase chain reaction) requiring hospitalization. Patients were followed up until intubation or exit. Patients that died in-hospital were excluded from primary analysis but were included in a multinomial model. Recorded data included days since diagnosis and symptom onset, type of oxygen delivery including non-invasive mechanical ventilation (NIMV), time from hospitalization onset to intubation or hospital discharge, radiological findings, pharmacological interventions (previous and disease related) ABGs analyses on admission and before intubation or hospital discharge. All arterial blood gases were analyzed on the same analyzer (ABL800 Basic©, Radiometer Medical ApS©).

### Outcomes

This study aimed to assess the relationship between hypocapnia (defined as pCO2 lower than 35 mmHg) upon admission and need for IMV in patients with COVID-19 pneumonia. Secondary outcomes included the impact of pCO2 on the composite outcome of intubation or mortality, the impact of NIMV on intubation and the combined effect of days since symptom onset and pCO2 upon admission on the incidence of intubation.

### Study size

Based on Riley et al.^13^, assuming a prevalence of 3.2% for intubation based on up-to-date published data^14^ and an R-squared of 18% for a model evaluating simultaneously up to three parameters (package pmsampsize for R, v. 4.0.2) a sample of 216 patients is required to draw safe conclusion with at least 7 events.

### Statistical methods

Appropriate descriptives were derived for all recorded variables. Logistic regression was used to assess the impact of pCO2 on the incidence of intubation. The predefined parameters (as defined in the study protocol) were assessed with univariate analysis: days since symptom onset and diagnosis, radiological severity, pO2 before intubation, type of NIMV prior to intubation. Odds ratios (OR) with 95% confidence intervals (CI) were calculated and reported along with the corresponding degrees of freedom. Taking into consideration sample characteristics, clinical significance and the univariate regression results, a multivariate model was built. A multinomial logistic regression model was fitted to incorporate data from deceased patients who were not intubated^15^. Data were recorded in an Access database. Analyses were run in R, v. 4.0.2. Missing data were not imputed, and incomplete cases were excluded from the respective analyses. No loss to follow-up occurred.

## RESULTS

### Characteristics of study population

Between October 2020 and January 2021, a total of 306 patients were hospitalized for COVID-19 pneumonia of variable severity. A total of 274 were included in the primary analysis ***(Fig.1)***; 6.9% (n=19/274) of them were intubated and 93% (n=255/274) were discharged. Baseline characteristics of the two groups are depicted in *Table 1*. No systematic differences were found for gender, age, history of pulmonary disease, prior anticoagulant therapy or comorbidities. Only 21% (n=4/19) of the intubated patients did not receive NIMV prior to intubation; all of them were deteriorating rapidly and immediate intubation and IMV was decided. Median time since symptom onset was 4 days (interquartile range-IQR: 6) in the discharge group and 3 days (IQR: 6) in the intubation group; median time between admission and intubation was 4 days (IQR: 6). Peak fraction of inspired oxygen (FiO2) needs was observed within the first three days from admission in 73.2% (n=201/274) of all patients.

**Table 1.** Patients demographics and past medical history

| **Characteristic** | **Non-intubated**<br>**patients**<br>**(N = 255)** | **Intubated**<br>**patients**<br>**(N = 19)** | **p-value<sup>1</sup>** |
| --- | --- | --- | --- |
| **Gender, n / N (%)** |  |  | **0.3** |
| Female | 110 / 255<br>(43%) | 6 / 19 (32%) |  |
| Male | 145 / 255<br>(57%) | 13 / 19 (68%) |  |
| **Age, Median (IQR)** | 64 (24) | 73 (15) | **0.11** |
| **History of pulmonary disease, n /**<br>**N (%)** |  |  | **0.086** |
| No | 224 / 255<br>(88%) | 14 / 19 (74%) |  |
| Yes | 31 / 255 (12%) | 5 / 19 (26%) |  |
| **Prior anticoagulant therapy, n / N**<br>**(%)** |  |  | **0.010** |
| No | 231 / 255<br>(91%) | 13 / 19 (68%) |  |
| Yes | 24 / 255<br>(9.4%) | 6 / 19 (32%) |  |
| **Days since symptom onset,**<br>**Median (IQR)** | 4.0 (6.0) | 3.0 (6.0) | **0.2** |
| **Days until intubation, Median**<br>**(IQR)** | NA (NA) | 4 (6) |  |
| Unknown | 255 | 0 |  |
| **NIMV, n / N (%)** |  |  | **<0.001** |
| No | 249 / 254<br>(98%) | 4 / 19 (21%) |  |
| Yes | 5 / 254 (2.0%) | 15 / 19 (79%) |  |
| Unknown | 1 | 0 |  |

| Characteristic | Non-intubated patients<br>(N = 255) | Intubated patients<br>(N = 19) | p-value<sup>1</sup> |
| --- | --- | --- | --- |
| **Type of NIMV, n / N (%)** |  |  | **<0.001** |
| BiPAP | 2 / 255 (0.8%) | 4 / 19 (21%) |  |
| CPAP | 1 / 255 (0.4%) | 2 / 19 (11%) |  |
| HFNC | 2 / 255 (0.8%) | 9 / 19 (47%) |  |
| NA | 250 / 255<br>(98%) | 4 / 19 (21%) |  |

**Abbreviations:** N: number; pts: patients; IQR: Interquartile range; NIMV: Non-invasive mechanical ventilation; BiPAP: bilevel positive airway pressure machine; CPAP: continuous positive airway pressure machine; HFNC: high flow nasal cannula; NA: not available.
<sup>1</sup>Pearson's Chi-squared test; Wilcoxon rank sum test; Fisher's exact test

**Figure 1.**
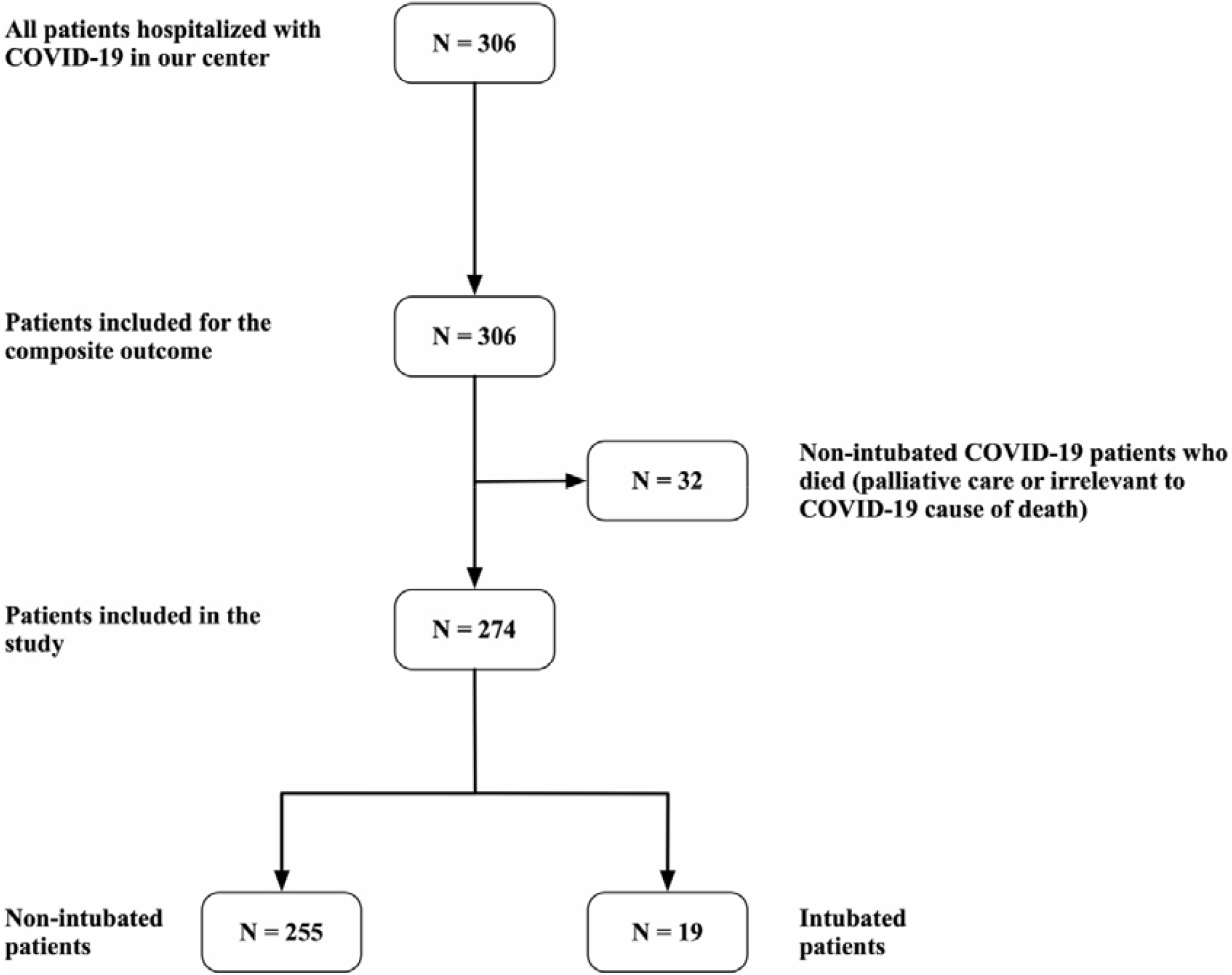

There was a statistically significant difference in baseline pCO2 in ABGs sample between the two groups; pCO2 was 33.6mmHg (median) in the discharge group (IQR: 5.7) and 29.7mmHg (IQR: 5.0) on the intubation group (Wilcoxon rank-sum test p.value = 0.003). Admission ABGs demonstrated a significant difference on pO2 and FiO2: median pO2 was 86mmHg (IQR: 34) in the discharge group and 69mmHg (IQR: 37) on the intubation group (Wilcoxon rank-sum test p.value = 0.020). Admission FiO2 needs were 21% (IQR: 11) in the discharge group and 44% (IQR: 59) in the intubation group (Wilcoxon rank-sum test p.value = 0.003). Follow-up ABGs analysis showed a similar trend for pO2 and FiO2, as median pO2 was 86mmHg (IQR: 22) in the discharge group and 52mmHg (IQR: 17) on the intubation group (Wilcoxon rank-sum test p.value < 0.001). Follow up FiO2 needs were 21% (IQR: 0) in the discharge group and 95% (IQR: 5) in the intubation group (Wilcoxon rank-sum test p.value < 0.001). There was no significant difference in follow up pCO2 (median 34.4mmHg (IQR: 5.9) in the discharge group and 32.2mmHg (IQR: 12.6) in the intubation group, Wilcoxon rank-sum test p.value = 0.8). There was no difference in total severity score between groups; however, no formal assessment for agreement was undertaken in the presented setting. Values of baseline measurements of ABGs analyses are presented in *Table 2*.

**Table 2.**
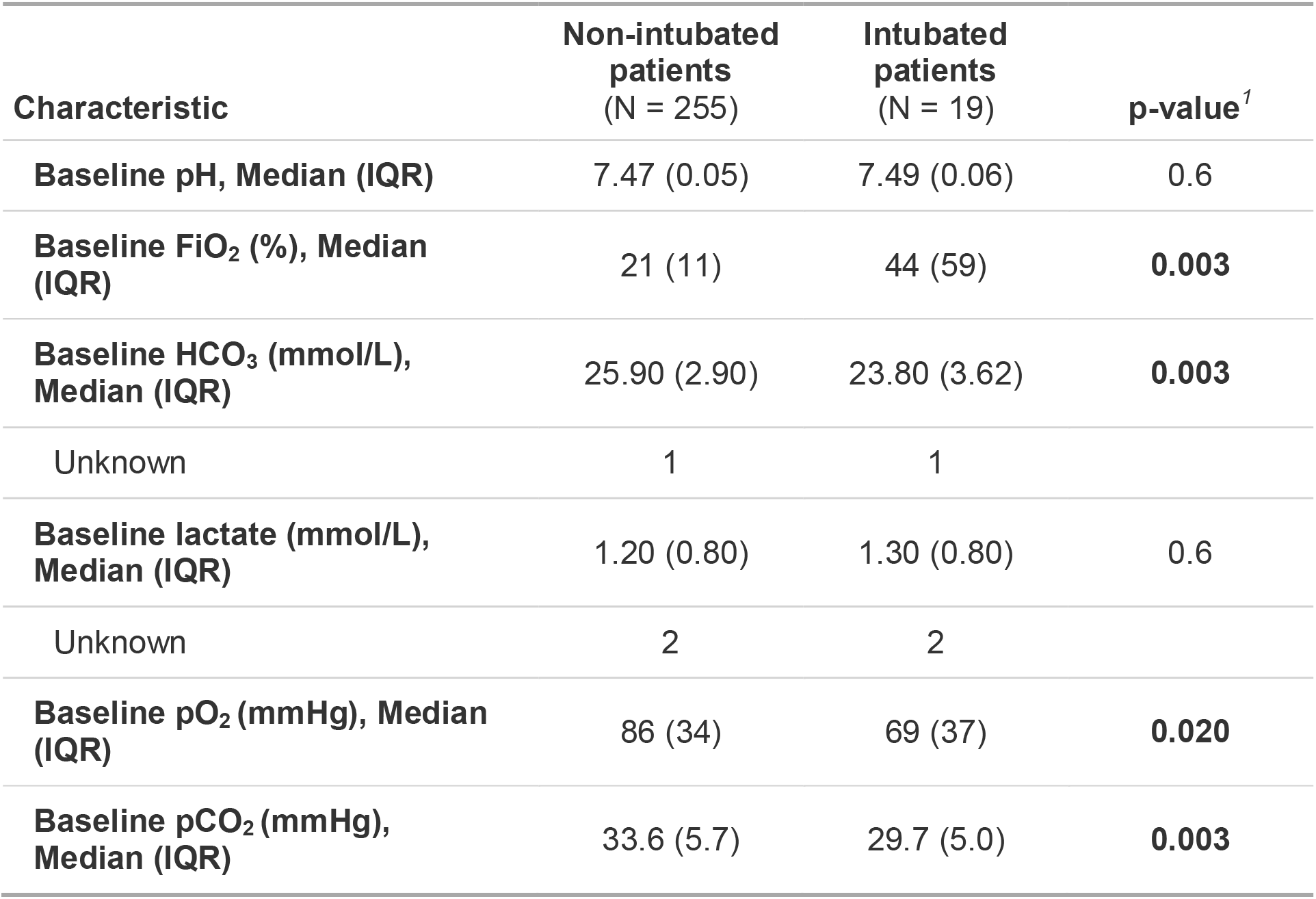
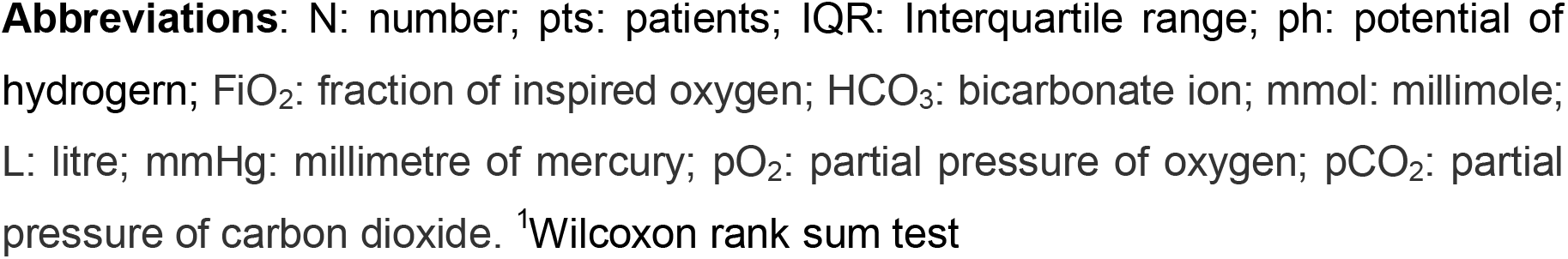
Values of baseline arterial blood gas analyses.

| Characteristic | Non-intubated patients<br>(N = 255) | Intubated patients<br>(N = 19) | p-value<sup>1</sup> |
| --- | --- | --- | --- |
| **Baseline pH, Median (IQR)** | 7.47 (0.05) | 7.49 (0.06) | 0.6 |
| **Baseline FiO<sub>2</sub> (%), Median (IQR)** | 21 (11) | 44 (59) | **0.003** |
| **Baseline HCO<sub>3</sub> (mmol/L), Median (IQR)** | 25.90 (2.90) | 23.80 (3.62) | **0.003** |
| Unknown | 1 | 1 |  |
| **Baseline lactate (mmol/L), Median (IQR)** | 1.20 (0.80) | 1.30 (0.80) | 0.6 |
| Unknown | 2 | 2 |  |
| **Baseline pO<sub>2</sub> (mmHg), Median (IQR)** | 86 (34) | 69 (37) | **0.020** |
| **Baseline pCO<sub>2</sub> (mmHg), Median (IQR)** | 33.6 (5.7) | 29.7 (5.0) | **0.003** |

**Abbreviations:** N: number; pts: patients; IQR: Interquartile range; ph: potential of hydrogen; FiO<sub>2</sub>: fraction of inspired oxygen; HCO<sub>3</sub>: bicarbonate ion; mmol: millimole; L: litre; mmHg: millimetre of mercury; pO<sub>2</sub>: partial pressure of oxygen; pCO<sub>2</sub>: partial pressure of carbon dioxide. <sup>1</sup>Wilcoxon rank sum test

Univariate analysis with logistic regression on the incidence of intubation is shown in *Table 3*. There was no statistically significant effect on the odds for intubation with respect to gender (OR: 1.64, 95% CI: 0.63, 4.80); age (OR: 1.03, 95% CI: 0.99, 1.06); history of pulmonary disease (OR: 2.58, 95% CI: 0.79, 7.28); days since symptom onset (OR: 0.92, 95% CI: 0.78, 1.05); use of NIMV (CPAP vs BiPAP: 1.00, 95% CI: 0.05, 29.8, HFNC vs BiPAP OR: 2.25, 95% CI: 0.21, 25.2) or time elapsed (in days) from symptom onset to the first available ABG sample (OR: 0.95, 95% CI: 0.89, 0.99). Baseline pO2 demonstrated borderline effect (OR: 0.98, 95% CI: 0.96, 1.00) with no useful clinical interpretation.

**Table 3.**
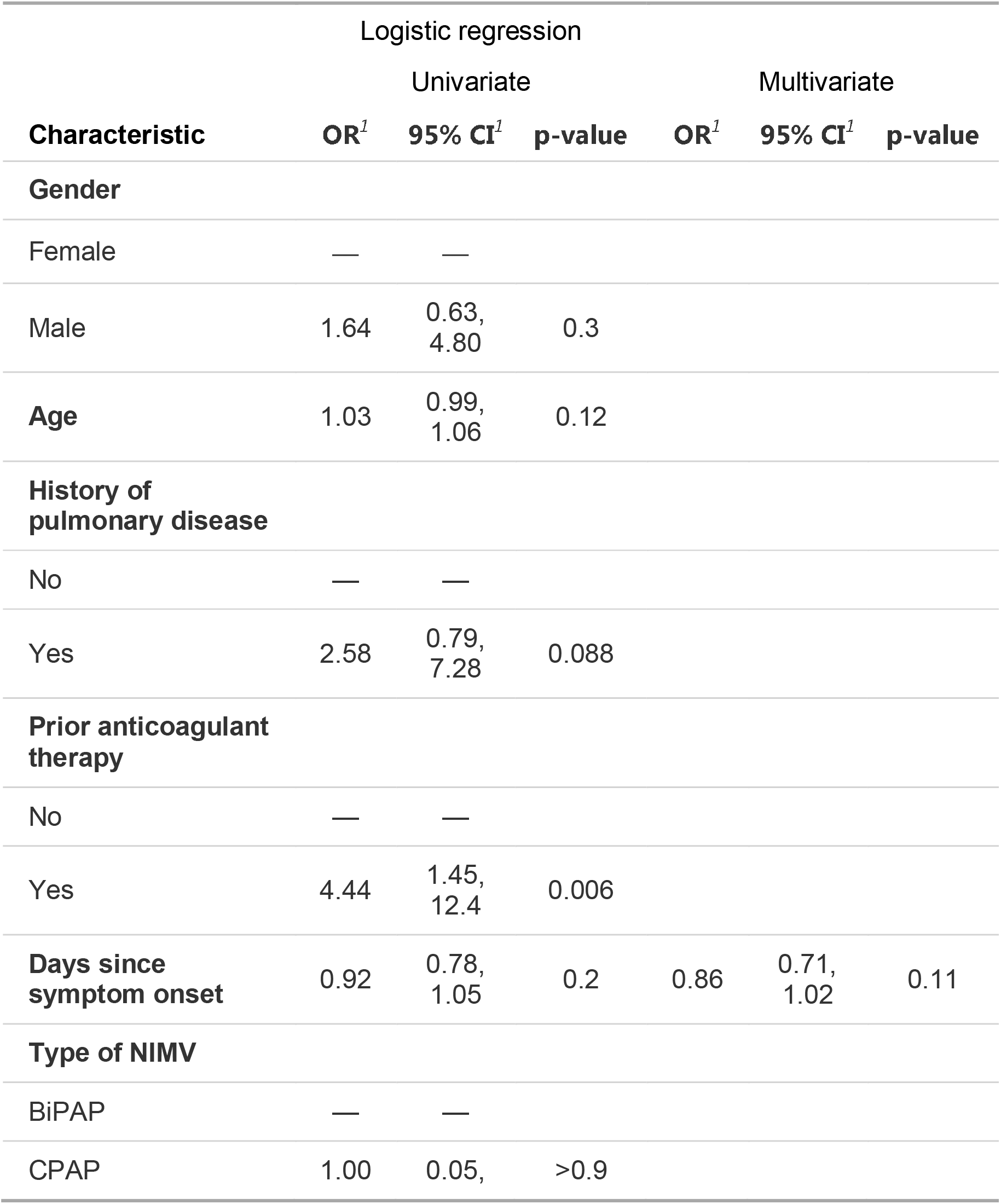
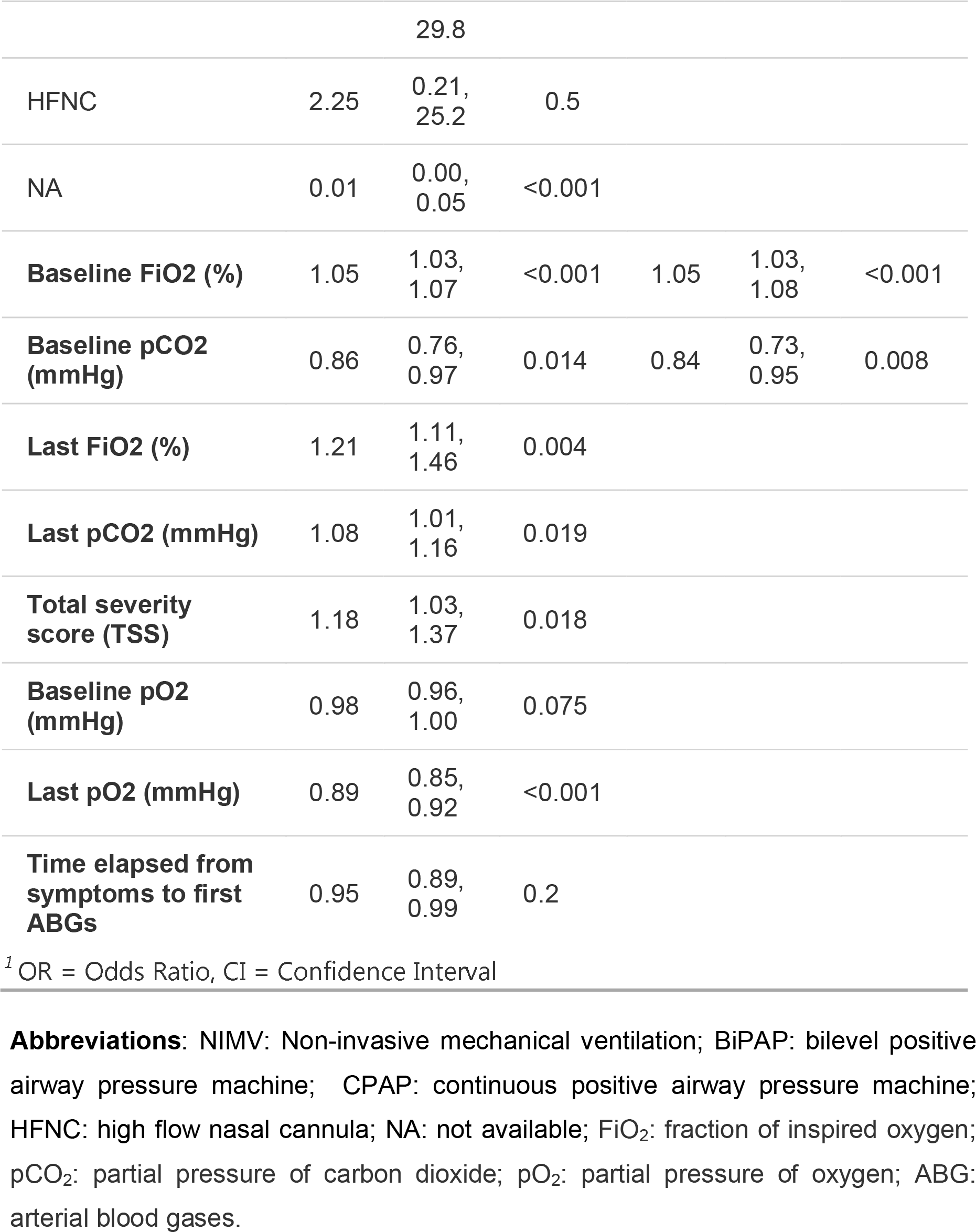
Results from statistical analysis

Parameters with statistically significant impact on the probability of intubation were baseline pCO2 measurement (OR: 0.86, 95% CI: 0.76, 0.97) and last pCO2 measurement before intubation (OR: 1.08, 95% CI: 1.01, 1.16), baseline FiO2 (OR: 1.05, 95% CI: 1.03, 1.07) as well as last FiO2 (OR: 1.21, 95% CI: 1.11, 1.46), total severity score on admission (OR: 1.18, 95% CI: 1.03, 1.37) and last pO2 (OR: 0.89, 95% CI: 0.85, 0.92). The primary outcome, the impact of baseline pCO2 on the odds for intubation, was found to increase by 16% for each unit of lower pCO2 on admission.

Taking into consideration sample characteristics, clinical significance and the univariate regression results, a multivariate model including baseline pCO2, days since symptom onset and baseline FiO2 was built *(Table 3)*. The odds for intubation increased with the decrease of pCO2 by 19% given the time since symptom onset and baseline FiO2 needs. A multinomial logistic regression incorporating data including deceased patients (deceased prior to intubation) was fitted. Intubation is still negatively associated with pCO2 levels (OR: 0.88, 95% CI: 0.78, 0.98); mortality was not significantly related to the pCO2 (OR: 1.06, 95% CI: 1.00, 1.13).

## DISCUSSION

In the current study we aimed to explore the relationship between a respiratory parameter, pCO2, in COVID-19 pneumonia and the risk for intubation. Our findings suggest an increased risk for intubation in those patients presenting with reduced pCO2, augmented by higher oxygen requirements and the advance of time since symptom onset.

To our knowledge, no study aiming to investigate this relationship has yet been designed. It is acknowledged that oxygen saturation alone should reflect the respiratory performance in patients with COVID-19 pneumonia being evaluated for intubation^16^. Published literature focuses on examining oxygen saturation and chemokines to determine those in danger for developing severe disease. However, from a physiological point of view, the evaluation of partial pressures measured in arterial blood is fundamental to associate prognosis and disease development and outcome.

Supporting evidence has been demonstrated in the literature published during the pandemic. Hypocapnia has been associated with worse outcome in a study by Brouqui et al.^16^ in the context of asymptomatic hypoxia in a sample of COVID-19 patients undergoing at least one ABGs analysis. Although the population under study was diagnosed with more severe disease and had a longer time since symptom onset, hypoxia – hypocapnia syndrome was strongly associated with intensive care unit admission and death whereas hypoxia – hypercapnia was associated with positive outcomes. Another study by Turcato et al.^17^ demonstrated a negative correlation of pCO2 and the extent of pulmonary invasion in computed tomography as well as mortality. Both study groups propose the further exploration of pCO2 as a predictive parameter.

A recently published multicenter prospective cohort study described that patients with ARDS due to COVID-19 infection have more thrombotic events, as they mainly develop coagulopathy associated with poorer prognosis^18^ compared to patients with non-COVID-19 ARDS^19^. The main thrombotic complication of these patients was pulmonary embolism causing severe hypoxia, hypocapnia and eventually respiratory distress^19^ while anticoagulant therapy might be associated with improved outcomes in patients with severe COVID-19^20^.

Several limitations are present in this observational study. The relatively small number of events, despite fulfilling the a priori power calculation to detect a difference, does not allow for generalization as a prognostic factor for intubation. This was a single centre study, with a limited number of cases; therefore caution is necessary when attempting to generalize the results. Future multicenter clinical trials included larger number of subjects are awaited to provide data about this prognostic factor. Another important constraint was the patient inflow, as the major proportion of patients were transferred from other hospitals, days after symptom onset and various stages of disease, having received various therapeutic agents. The baseline measurements of our study may not be homogenous with respect to days since symptom onset and treatment naïveté or nosocomial infection; in a larger sample this heterogeneity would yield a much more robust result. Another important constraint is the lack of evaluation for observer agreement for the radiological findings of chest computed tomography. Our data was not sufficient to run such analyses; as a result, this important parameter of disease severity could not be reliably assessed as a contributing factor.

Despite the limitations, there are several strengths in the current comparison. The most important is the inclusion of all admitted patients, regardless of comorbidities or previous respiratory disease. This allows for generalization of our main conclusion to a broader group of patients requiring hospitalization for COVID-19 pneumonia. Additionally, the use of a composite outcome allowed for inclusion of all patients, eliminating selection bias that would occur when excluding deceased patients. In the last year, numerous prognostic models for COVID-19 pneumonia have been published aiming to recognize as early as possible those patients who will require intensive care during their treatment. Most of them, despite reporting high C scores, are not externally validated or are deemed of high risk of bias, therefore their clinical applicability is limited^21^. Adding pCO2 as a prognostic factor might be of clinical significance in these models and should be further investigated.

## CONCLUSION

The dissociation between respiratory failure and a clinically comfortable patient is partly due to decreased carbon dioxide levels associated with COVID-19 pneumonia. Hypocapnia seems to be a determinant factor for intubation in patients with COVID-19 pneumonia in this study.

## Supporting information

STROBE checklist

## Data Availability

All data produced in the present study are available upon reasonable request to the authors.

## FUNDING

There were no funding sources for the current study

## CONFLICT OF INTEREST

All contributing authors have nothing to declare.

## Notes

### Competing Interest Statement

The authors have declared no competing interest.

### Funding Statement

This study did not receive any funding.

### Author Declarations

Ethics and scientific conduct committee of Proto Geniko Nosokomeio Thessalonikis O Agios Pavlos gave ethical approval for this work.

